# Association between depressive symptoms and physical function among participants with heart disease in the Reasons for Geographic And Racial Differences in Stroke (REGARDS) study

**DOI:** 10.64898/2026.06.09.26355319

**Authors:** Mojisola E. Fasokun, Monika M. Safford, Yulia Khodneva, Lisandro D. Colantonio, Parag Goyal, Chibuike J Alanaeme, Abu Abdullah Mohammad Hanif, Ene M. Enogela, C. Barrett Bowling, Emily B. Levitan

**Affiliations:** Department of Epidemiology, University of Alabama at Birmingham, Birmingham, AL; Division of General Internal Medicine, Weill Cornell Medicine, New York, NY; Department of Medicine School of Medicine University of Alabama at Birmingham AL; Department of Medicine, Duke University School of Medicine, Durham NC; Durham Veterans Affairs Geriatric Research Education and Clinical Center, Durham Veterans Affairs Medical Center (VAMC), Durham, NC

**Author notes:** Corresponding author: Emily B. Levitan, Professor and Vice Chair for Faculty Development Department of Epidemiology University of Alabama at Birmingham.

**Keywords:** Depressive symptoms, Heart disease, Physical function

## Abstract

**Background:** Depression and heart disease frequently co-occur in the aging population and are associated with functional decline and poor health outcomes. Understanding how depressive symptoms relate to different aspects of physical function among adults with heart disease may help identify high-risk subgroups.

**Objective:** To examine the association of depressive symptoms with self-reported and observed physical function measures among participants with heart disease in the Reasons for Geographic and Racial Differences in Stroke (REGARDS) study and assess whether associations differ by sex and race–sex groups.

**Methods:** We conducted a cross-sectional analysis using data from REGARDS study second in-home visit (2013–2016). Depressive symptoms were measured with the 10-item Center for Epidemiologic Studies Depression scale (CES-D-10), considering scores ≥10 as clinically significant. Physical function measures were instrumental activities of daily living (IADL), activities of daily living (ADL), chair stand time (5 repetitions), and gait speed. Linear regression models estimated associations of depressive symptoms with function, adjusting for sociodemographic, health behavior, antidepressant medications, body mass index, and social support. Effect modification by sex and race–sex group was evaluated.

**Results:** Among 3,055 participants, 11.7% had CES-D-10 ≥10. Compared to CES-D-10 scores <10, CES-D-10 ≥10 was associated with more limitations in IADL (1.84 points; 95% CI 1.62, 2.06), ADL (0.43 points; 95% CI 0.34, 0.52) and slower chair stand time (0.88 second; 95% CI 0.07, 1.69); associations with gait speed were modest (−0.04 meters/second; 95% CI −0.08, - 0.01). Women had a stronger association between CES-D-10 and ADL (0.49 points; 95% CI 0.35, 0.64) than men (0.33 points; 95% CI 0.21, 0.44; p for interaction = 0.01). Interaction between CES-D-10 and race–sex groups was not statistically significant.

**Conclusions:** Among adults with heart disease, clinically significant depressive symptoms were associated with lower physical function, particularly among women.

## Introduction

Cardiovascular disease (CVD) and depression are major public health challenges in the United States, as each significantly impacts the adult population, and they both contribute to mortality, morbidity, disability, hospitalization and economic burden^1–4^. Depressive symptoms influence recovery and quality of life (QoL) after myocardial infarction^5^ and increases the risk of future cardiac events or death, regardless of CVD severity^6^.

However, the impact of depressive symptoms on physical function in heart disease is not well understood, even though physical function is a key patient-centered outcome in CVD care^7,8^. Physical function, which reflects a person’s ability to perform daily activities and remain independent^9^, is strongly linked to outcomes including hospitalization, nursing home admission, falls, and death^10^. Functional limitation is common in individuals with both CVD and depression, contributing to disability and reduced QoL^11,12^.

Prior research shows mixed results for differences in associations of mental health and mental health interventions by sex, with outcomes including clinical event rates, psychosocial improvements, or recovery trajectories^13,14^. Examining sex and race jointly offers deeper insight into health outcomes and disparities, especially for groups like Black women, who experience a disproportionate burden and worse outcomes relative to other subgroups^15–17^. This study aims to examine the association between depressive symptoms and physical function markers among participants with heart disease in the Reasons for Geographic and Racial Differences in Stroke (REGARDS) study, overall and by sex and race–sex subgroups.

## Methods

### Study Population

The REGARDS study is a large, national, community-based cohort that enrolled 30,239 self-identified Black or White adults aged 45 years or older from across the contiguous US states (2003–2007), with oversampling of Black individuals and residents of the Southeastern "Stroke Belt/Buckle". At baseline, participants completed a computer-assisted telephone interview to collect information on sociodemographics, health behaviors, and medical history, followed by an in-home visit during which trained technicians measured blood pressure, anthropometrics, electrocardiograms, and collected blood and urine samples. Participants have been followed every six months by telephone for updates on hospitalizations, stroke and other cardiovascular outcomes, and general health status. Approximately 10 years after baseline, a second round of telephone interviews and in-home visits was conducted (2013–2016) to update health status and collect additional data, including expanded assessments of physical function^18,19^.

For this study, we focused on individuals with a history of heart-related CVD present by the time of their second in-home visit, including self-reported or adjudicated coronary heart disease, heart failure (HF), and atrial fibrillation, and those who had undergone cardiac revascularization procedures. Participants with a history of stroke were excluded to minimize heterogeneity, as stroke can significantly impair physical function through neurologic mechanisms distinct from other CVDs, thus the association between depressive symptoms and physical function may be different among individuals with a history of stroke^20^. Individuals were excluded if they lacked data on either depressive symptoms or physical function assessments (**Supplementary Figure 1**).

#### Depressive symptoms

Depressive symptoms were measured using the Center for Epidemiologic Studies Depression Scale –10 item version (CES-D-10). Items are summed to yield a total score ranging from 0 to 30, with higher scores indicating more severe depressive symptoms^21^. A score ≥10 for the 10-item version was considered the cutoff for screening for clinically significant depressive symptoms, consistent with established, validated scoring guidelines^22^.

### Self-reported and Observed Physical Function

#### Instrumental Activities of Daily Living (IADL) Measurement

The IADL assessment evaluated participants’ self-reported ability to carry out complex daily tasks that required both cognitive and physical functions (e.g., managing finances, taking medications). Responses are categorized as follows: 0 = "I could do it by myself with no difficulty"; 1 = "I could do it by myself with some difficulty"; and 2 = "I would need someone to help me". The scores for each IADL activity were summed, yielding a total score ranging from 0 to 14^23^.

#### Activities of Daily Living (ADL) Measurement

ADL measurement similarly involved participants self-reporting their ability to perform basic personal care tasks (e.g., getting out of bed or chair, dressing). The ADL scores are calculated across the activities, with an overall score ranging from 0 to 10^23^.

Although a formal minimal clinically important difference has not been established for IADL and ADL summary scores, a 1-point higher score reflects one additional daily activity performed with difficulty or requiring help, representing a meaningful functional change^24^.

#### Chair Stand Test

Participants were asked to complete 5 chair stands, completely standing up and sitting down without using their arms. The time to complete these 5 chair stands was recorded, with times outside the 1-second to 1-minute range considered invalid. The chair stand test was used as an indicator of lower body strength and balance, with longer times to complete indicating worse function^25^.

#### Gait Speed Test

Participants performed two 8-foot timed walks, with the option to use canes or walkers if needed. The average time of these walks was converted to gait speed in meters per second. Times greater than 1 minute or less than 1 second were considered invalid^25^.

Small but meaningful changes in gait speed are on the order of 0.05 m/s, with larger clinically important changes around 0.10 m/s, and a minimal clinically important difference of about 2–3 seconds has been reported for the 5-times chair stand test^26,27^.

#### Covariates

Covariates were selected based on established or plausible associations between depressive symptoms and physical function, as reported in prior literature^28^. These included age, sex, race, education level, income level, relationship status, self-reported alcohol use, smoking status, antidepressant medications^28,29^, body mass index (BMI, kg/m²), and social support measured using the ENRICHD Social Support Instrument (ESSI), with low support defined as ESSI total score ≤ 18^30^. Diabetes, hypertension, and chronic kidney disease (CKD) were summarized descriptively but excluded from primary models to avoid overadjustment bias for comorbidities potentially on the causal pathway.

Race, as a social construct, was included as confounder and potential effect modifier because it serves as a proxy for social determinants of health that may influence both depressive symptoms and physical function and because prior studies have demonstrated differences in the psychometric properties of the CES-D across racial and ethnic groups ^31–34^.

### Statistical analysis

We conducted descriptive statistical analyses to summarize the characteristics of study participants. For the primary analyses, we employed linear regression models to examine associations between depressive symptoms and each physical function marker, specifically IADL score, ADL score, chair stand time, and gait speed. To account for potential non-normality of residuals and address potential violations of model assumptions, empirical variance estimators were applied. Three models were specified: Model 1 was unadjusted; Model 2 was adjusted for age, sex, race, education, income, and relationship status; and Model 3 was further adjusted for alcohol use, smoking, and BMI, as well as social support as measured with ESSI, and antidepressant medications^35^. We also fitted models by adding each covariate individually to the unadjusted model to explore which variables contributed most to attenuation of the unadjusted association.

To assess whether sex modifies the associations between depressive symptoms and physical function, we included interaction terms in the regression models and interactions were considered statistically significant at a p-value < 0.05. We also evaluated whether the association between depressive symptoms and physical function varied across four race-sex groups (Black women, Black men, White women, and White men) by including interaction terms for race-sex group, and depressive symptoms. Missing covariate data were addressed using multiple imputation by chained equations.

For sensitivity analyses, we excluded participants with HF from the study population (**Supplementary Figure 2**) based on evidence suggesting that the relationship between depression and HF may differ from its relationship with other CVD subtypes^36^. We also repeated the main analyses using an alternative CES-D-10 cutoff (≥16)^37^ representing more severe depressive symptoms^38^. Physical function measures were also analyzed as dichotomous outcomes using logistic regression. IADL and ADL impairment were defined as reporting "I would need someone to help me" or “I could do it by myself with some difficulty”^19^ for one or more activities. For the chair stand test, participants who required more than 13.6 seconds or were unable to complete the test were classified as lower performance, while those completing the test in ≤13.6 seconds were considered higher performance consistent with prior work^39^. For gait speed, we applied a cutoff of <0.6 m/s analysis or not completing the test, consistent with previous studies showing that this threshold identifies individuals at high risk for physical disability and dependency^40,41^. As a result of including individuals who could not perform the gait speed and chair stand tests, the analytic sample for models based on these definitions was larger than the sample used for linear regression models of continuous outcomes (**Supplementary Figure 3**).

#### Ethical approval and consent

Ethical approval for the REGARDS study was obtained from the Institutional Review Boards (IRB) at the participating institutions. Written informed consent was obtained during the in-home visit.

## Result

Of the 3,055 participants, 356 (11.7%) had clinically significant depressive symptoms (CES-D-10 score ≥10) (**Table 1**). Compared to those with CES-D-10 <10, participants with scores ≥10 were younger (72.6 ± 8.1 vs. 74.8 ± 7.7 years), and a greater proportion was female (62.1% vs. 37.2%) and Black (36.8% vs. 27.9%), a smaller proportion had a college education (22.8% vs. 43.0%) and a larger proportion had annual incomes < $20,000 (25.3% vs. 11.1%). Clinical comorbidities were more common in the CES-D-10 ≥10 group, including BMI ≥30 (50.8% vs. 37.1%), diabetes (42.4% vs. 30.9%), and hypertension (84.0% vs. 79.6%), while CKD prevalence was similar (36.3% vs. 34.5%). Participants with CES-D-10 ≥10 had lower alcohol use (29.2% vs. 44.4%), social support (ESSI score 25.0 ± 6.4 vs. 28.8 ± 5.3) and higher smoking (12.7% vs. 7.0%) and antidepressant use (51.4% vs. 13.5%).

**Table 1.** Characteristics of REGARDS participants with heart disease by clinically significant depressive symptoms.

|  | **Overall**<br>**(N= 3,055)** | **CES-D-10 score <10**<br>**(n= 2,699)** | **CES-D-10 score ≥10**<br>**(n= 356)** |
| --- | --- | --- | --- |
| **Sociodemographic** |  |  |  |
| **Age, years (mean ± SD)** | 74.6 ± 7.8 | 74.8 ± 7.7 | 72.6 ± 8.1 |
| **Sex, n (%)** |  |  |  |
| Female | 1225 (40.1) | 1004 (37.2) | 221 (62.1) |
| Male | 1830 (59.9) | 1695 (62.8) | 135 (37.9) |
| **Race, n (%)** |  |  |  |
| Black | 884 (28.9) | 753 (27.9) | 131 (36.8) |
| White | 2171 (71.1) | 1946 (72.1) | 225 (63.2) |
| **Education, n (%)** |  |  |  |
| < High school | 235 (7.7) | 183 (6.8) | 52 (14.6) |
| High school graduate | 770 (25.2) | 655 (24.3) | 115 (32.3) |
| Some college | 808 (26.5) | 700 (25.9) | 108 (30.3) |
| College graduate & above | 1242 (40.7) | 1161 (43.0) | 81 (22.8) |
| **Income, n (%)** |  |  |  |
| <\$20,000 | 390 (12.8) | 300 (11.1) | 90 (25.3) |
| \$20,000-\$34,000 | 744 (24.4) | 649 (24.0) | 95 (26.7) |
| \$35,000-\$75,000 | 971 (31.8) | 884 (32.8) | 87 (24.4) |
| \$75,000 and above | 558 (18.3) | 534 (19.8) | 24 (6.7) |
| Declined to report | 392 (12.8) | 332 (12.3) | 60 (16.9) |
| **Relationship Status, n (%)** |  |  |  |
| Married | 1855 (60.7) | 1685 (62.4) | 170 (47.8) |
| **Health Status** |  |  |  |
| **BMI (kg/m<sup>2</sup>), n (%)** |  |  |  |
| <25 | 629 (20.6) | 575 (21.3) | 54 (15.2) |
| 25 - 29.9 | 1244 (40.7) | 1123 (41.6) | 121 (34.0) |
| ≥30 | 1182 (38.7) | 1001 (37.1) | 181 (50.8) |
| Diabetes, n (%) | 956 (32.2) | 809 (30.9) | 147 (42.4) |
| Hypertension, n (%) | 2446 (80.1) | 2147 (79.6) | 299 (84.0) |
| History of CKD, n (%) | 1014 (34.7) | 892 (34.5) | 122 (36.3) |
| **Lifestyle** |  |  |  |
| **Any Alcohol use, n (%)** | 1297 (42.7) | 1194 (44.4) | 103 (29.2) |
| **Cigarette smoking, n (%)** |  |  |  |
| Current | 233 (7.7) | 188 (7.0) | 45 (12.7) |
| Never | 1306 (43.0) | 1151 (42.9) | 155 (43.8) |
| Past | 1498 (49.3) | 1344 (50.1) | 154 (43.5) |
| **Psychosocial Factors (Social Support), (mean ± SD)** |  |  |  |
| ESSI score | 28.4 ± 5.5 | 28.8 ± 5.3 | 25.0 ± 6.4 |
| **Medications, n (%)** |  |  |  |
| Antidepressant | 544 (17.9) | 362 (13.5) | 182 (51.4) |

**Abbreviation:** BMI = Body Mass Index; CKD = chronic kidney disease, ESSI = ENRICHD Social Support Instrument
CES-D-10 score $\geq 10$ indicates clinically significant depressive symptoms; CES-D-10 score $< 10$ indicates non-clinically significant depressive symptoms.
Body Mass Index (BMI, kg/m<sup>2</sup>) grouped as normal/underweight ( $< 25$ ), overweight (25–29.9), and obese ( $\geq 30$ )

### Associations between Clinically Significant Depressive Symptoms and Physical Function

Compared to CES-D-10 <10, participants with CES-D-10 ≥10 had higher IADL scores (2.15 points; 95% CI 1.82, 2.49, unadjusted), which remained significant after partial and full adjustment (**Figure 1, Supplementary Table 1**). ADL scores were also higher (0.52 points; 95% CI 0.38, 0.67, unadjusted) and remained significant across all models. Participants with CES-D-10 ≥10 had slower chair-stand performance in unadjusted models (mean difference 2.08 seconds; 95% CI 1.20, 2.96), which was attenuated in partial and fully adjusted models. For gait speed, the unadjusted mean difference was −0.08 meters/second (95% CI −0.11, −0.05), which was attenuated with partial adjustment and remained significant in the fully adjusted model. For IADL, ADL, chair-stand, and gait speed, stepwise covariate adjustment attenuated the associations, with no single variable exerting a dominant effect (**Supplementary Table 2**). After excluding participants with HF, CES-D-10 had a similar pattern of results as observed in the main analyses (**Supplementary Table 3**).

**Figure 1.**
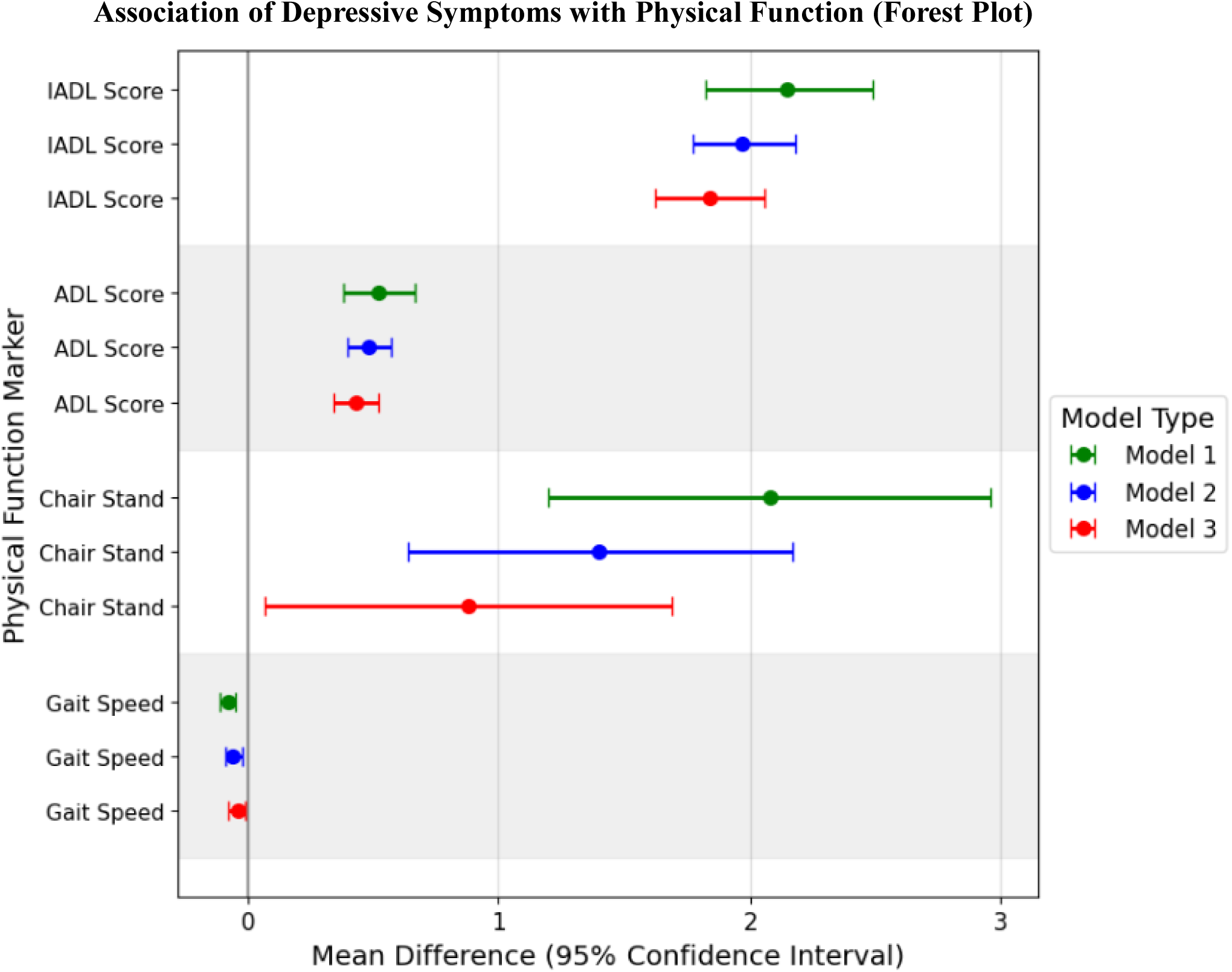
(Forest Plot): Associations between clinically significant depressive symptoms and physical function markers among REGARDS study participants with heart disease. Regression coefficients (β, in points for IADL and ADL, seconds for chair stand and meters/second for gait speed) and 95% confidence intervals (CI) represent the estimated difference in physical function markers between participants with and without clinically significant depressive symptoms (CES-D-10 score ≥10). The vertical line at zero represents no difference. Three models are presented: Model 1 (unadjusted); Model 2 (adjusted for sociodemographic factors); and Model 3 (Model 2 plus health behaviors, medication use, and ESSI social support).

### Associations Between Clinically Significant Depressive Symptoms and Physical Function Outcomes by Sex and Race-Sex group

Black women had the highest prevalence of CES-D-10 ≥ 10 and the greatest IADL and ADL scores, whereas White men had the lowest prevalence of CES-D-10 ≥ 10 and the best physical performance (**Supplemental Table 4**). For IADL, chair-stand, and gait speed, the interaction terms between sex and CES-D-10 ≥ 10 were not statistically significant. The interaction term for ADL was statistically significant in both the partially adjusted (p = 0.02) and fully adjusted models (p = 0.01); therefore, ADL models were stratified by sex. Among women, CES-D-10 ≥ 10 was more strongly associated with ADL score compared with men (unadjusted difference for women 0.58; 95% CI 0.38, 0.79; for men 0.38; 95% CI 0.18, 0.58), though the associations were attenuated after adjustment (**Table 2**). The overall interaction between depressive symptoms and race–sex group was not statistically significant for IADL, ADL, chair stand or gait speed, (p-value for overall interaction = 0.48, 0.35, 0.22, and 0.85, respectively).

**Table 2.** Associations between clinically significant depressive symptoms and ADL outcomes by sex among REGARDS study participants with heart disease.

|  | **Male** |  | **Female** |  |  |
| --- | --- | --- | --- | --- | --- |
|  | CES-D-10 score<br>≥10 | CES-D-10 score<br><10 | CES-D-10 score ≥10 | CES-D-10 score<br><10 | P-value |
| **ADL Score, points (possible range 0 –10, higher scores indicate worse function)** |  |  |  |  |  |
| **Model 1**<br>**Difference**<br>**(95% CI)** | 0.38 (0.18,0.58) | Reference | 0.58 (0.38,0.79) | Reference | 0.16 |
| **Model 2**<br>**Difference**<br>**(95% CI)** | 0.37 (0.27,0.49) | Reference | 0.55 (0.36,0.76) | Reference | 0.02 |
| **Model 3**<br>**Difference**<br>**(95% CI)** | 0.33 (0.21,0.44) | Reference | 0.49 (0.35,0.64) | Reference | 0.01 |

**Abbreviations:** ADL = Activities of Daily Living, CI = Confidence Interval Differences ( $\beta$ ) represent the mean difference in each physical function marker for participants with depressive symptoms compared to those without. Model 1: Unadjusted. Model 2: Model 1 + sociodemographic factors (age, sex, race, education, income, and relationship status). Model 3: further adjusted for health behavioral factors (alcohol use, smoking status), body mass index (BMI), medications (antidepressants) and social support using ENRICH Social Support Instrument (ESSI). P-values correspond to the interaction term.

### Sensitivity analysis

Compared to participants with CES-D-10 <16, those with CES-D-10 ≥ 16 (n = 172) had greater IADL scores across all models (2.91 points; 95% CI 2.24, 3.59, unadjusted), and ADL scores were consistently higher among participants with CES-D-10 ≥ 16 than among those with CES-D-10 < 16 (**Supplementary Table 5**). Those with CES-D-10 ≥ 16 had slower chair stand performance in the unadjusted model (3.30 seconds; 95% CI 1.52, 5.05), with associations attenuated after partial and full adjustment. For gait speed, the unadjusted mean difference was - 0.09 meters/second (95% CI -0.14, -0.04), which was attenuated and not statistically significant after full adjustment.

The interaction terms between CES-D-10 ≥ 16 and sex in both the partially and fully adjusted models were significant for IADL (p =0.01 and p < 0.01, respectively) and ADL (p =0.01 in both models) (**Supplementary Table 6**). Therefore, subsequent analyses for IADL and ADL were stratified by sex. Among women, CES-D-10 ≥ 16 was strongly associated with worse IADL and ADL compared with men, though the associations were attenuated after adjustment (fully adjusted IADL difference for women: 2.67; 95% CI 2.14, 3.20; for men: 1.89; 95% CI 1.37, 2.40 points). However, the interaction terms between sex and CES-D-10 ≥ 16 were not significant for chair stand or gait speed.

Using dichotomous outcomes (**Figure 2, Supplemental Table 7**), compared to participants with CES-D-10 <10, those with CES-D-10 ≥ 10 had significantly higher odds of having difficulty or needing help with at least one IADL and ADL, slow chair stand and slow gait speed and these associations were attenuated but remained statistically significant after adjustment. In all models, participants with CES-D-10 ≥ 10 had more than 4-fold higher odds of IADL impairment when compared with those CES-D-10 <10, (5.05; 95% CI 4.06, 6.30, unadjusted). CES-D-10 ≥ 10 was associated with 2-fold higher odds of slow chair stand (2.16; 95% CI 1.77, 2.66) and slow gait speed (2.39; 95% CI 1.73, 3.25) in unadjusted models compared to those with CES-D-10 <10.

**Figure 2.**
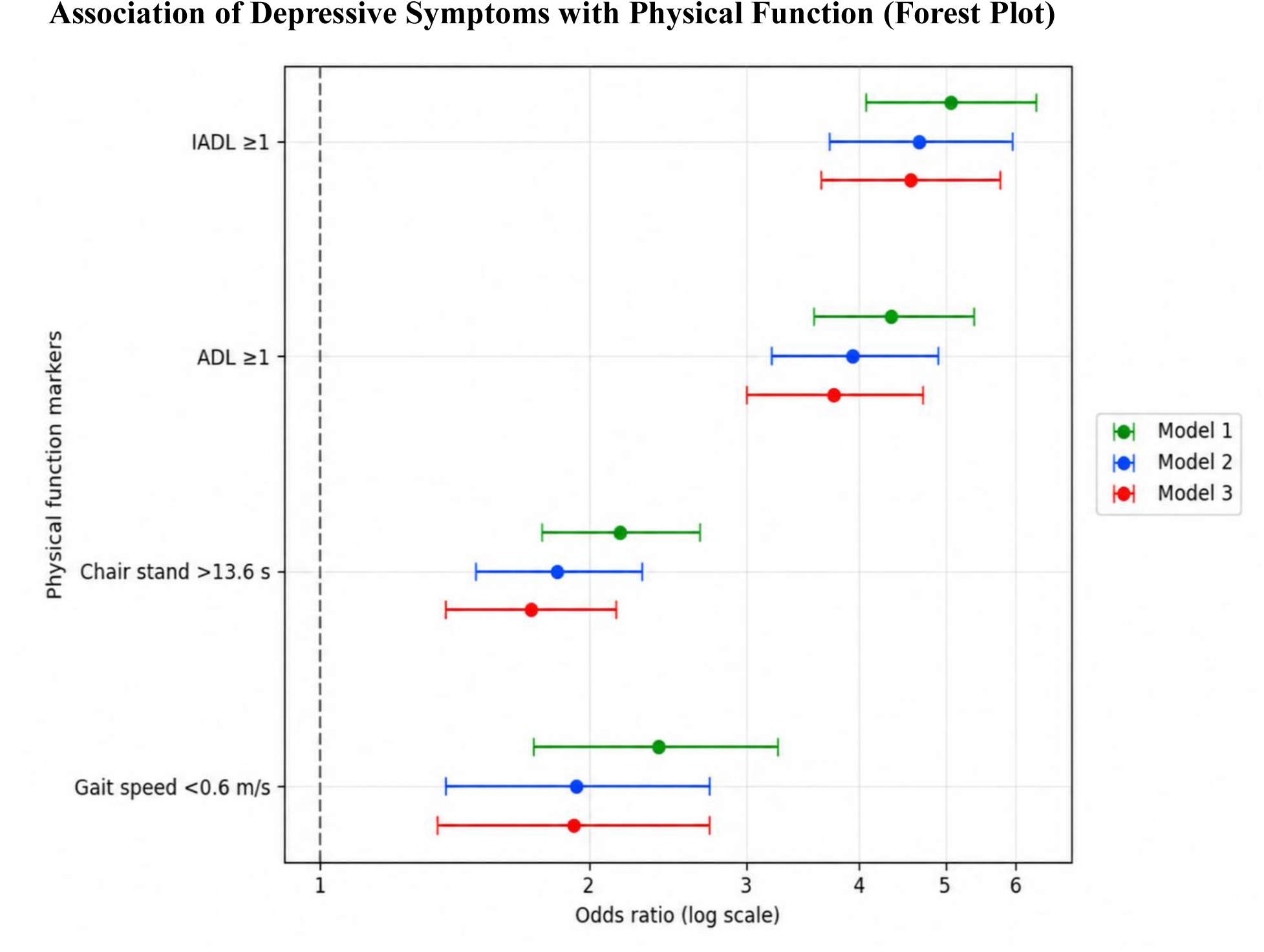
Associations between clinically significant depressive symptoms and physical function markers among REGARDS study participants with heart disease. Odds ratios and 95% confidence intervals are shown for the association between clinically significant depressive symptoms (CES-D-10 ≥10 vs <10) and each physical function marker. Model 1 is unadjusted; Model 2 is adjusted for age, sex, race, education, income, and relationship status; Model 3 is additionally adjusted for alcohol use, smoking status, body mass index, antidepressant use, and social support (ENRICHD Social Support Instrument). Analyses were conducted using logistic regression. Odds ratios are displayed on a log scale, and the dashed vertical line at OR = 1.0 indicates no association.

## Discussion

In this population of adults with history of heart disease, depressive symptoms were associated with worse self-reported and observed physical function, including greater IADL and ADL impairment and slower chair-stand and gait-speed performance in unadjusted models; this underscores the need to recognize potential function limitations in individuals with depressive symptoms. Depressive symptoms were associated with worse IADL and ADL scores, even after adjustment for covariates, with a suggestion that the associations were stronger among women than among men. In fully adjusted models, CES-D-10 ≥10 and ≥16 were associated with slower chair stand times, whereas the association with gait speed was significant only at CES-D-10 ≥10.

These results extend prior studies linking depression to functional impairment in older adults by focusing on individuals with established heart disease, a population at high risk of both adverse mental health and function limitations^42^. These findings suggest that depressive symptoms may be more strongly related to self-reported limitations in daily activities than observed lower-extremity performance in this population, consistent with previous studies^43^. This distinction is important, because patient-reported function is a key determinant of QoL, care needs, and prognosis in CVD^44^, and self-report and performance-based measures capture related but non-interchangeable aspects of physical functioning.

The persistence of associations after adjustment for demographic characteristics, health behaviors, social support, and antidepressant use supports the view that depression may contribute to functional vulnerability in heart disease, potentially via pathways such as reduced activity, adverse social environments, systemic inflammation, neuroendocrine dysregulation, or poor engagement in rehabilitation and self-care^45,46^, thereby increasing the risk of hospitalizations, nursing home admissions, falls, greater dependency, and death^46,47^. Routine assessment of depressive symptoms in CVD care may help identify individuals who warrant more detailed evaluation of functional status. In our sample, only about half of participants with clinically significant depressive symptoms were taking antidepressants, highlighting a substantial gap in depression care among CVD patients; data on non-pharmaceutical mental health therapies was not available. Our findings reinforce current recommendations that underscore the importance of treatment of depression as a key component of care for patients with CVD^48,49^.

Women showed a stronger association between CES-D-10 ≥10 and ADL outcomes, and at CES-D-10 ≥16, the associations were stronger for both ADL and IADL. These findings align with previous studies suggesting that women may experience a greater functional impact and may have different recovery trajectories and care experiences than men with CVD, potentially reflecting differences in symptom perception, care-seeking, social roles, and exposure to structural and interpersonal stressors^19,50^. These findings support prior work arguing for tailoring interventions to gender-specific psychosocial needs^51^. CES-D-10 and race–sex group interaction were not significant which may be due to low power for the test. An intersectional framework that considers how race, sex, socioeconomic status, and other structural factors interact may be necessary to understand how these characteristics jointly shape depression-related functional limitations^15–17,52^ and to design interventions.

## Strengths and Limitations

This study utilized a large, geographically diverse cohort, standardized self-reported assessment of depressive symptoms and validated performance-based physical function measures as well as self-reported function.

The study has several limitations. Depressive symptoms involves both psychological and physical manifestations, including reduced physical activity, creating a bidirectional relationship that complicates causal inference^53^. The cross-sectional study design further limits causal inferences due to potential reverse causality^54^. Residual confounding by unmeasured factors such as pain may persist despite adjustment. CES-D-10 is a screening tool rather than a diagnostic interview, and misclassification is possible. REGARDS include only Black and White adults ages 45 years and older, which may limits generalizability. Further, only community-dwelling adults were included, thus missing nursing home residents, who may have higher rates of depressive symptoms.

## Conclusion

Among adults with heart disease in the REGARDS study, clinically significant depressive symptoms were associated with worse self-reported and observed physical function, with a suggestion that the association with self-reported function was stronger among women. This provides additional evidence that addressing depression is an important component of patient-centered cardiovascular care.

## Data Availability

Data availability: The REGARDS study shares data with investigators through formal data use agreements. Requests for information on this process and data access may be sent to the REGARDS study at. Data availability statement: Data are available upon reasonable request. Data may be obtained from a third party and are not publicly available.

## Acknowledgement

The REGARDS study is supported by cooperative agreement U01 NS041588 co-funded by the National Institute of Neurological Disorders and Stroke (NINDS) and the National Institute on Aging (NIA), National Institutes of Health, Department of Health and Human Service. The content is solely the responsibility of the authors and does not necessarily represent the official views of the NINDS or the NIA. Representatives of the NINDS were involved in the review of the manuscript but were not directly involved in the collection, management, analysis or interpretation of the data. Additional funding for this work was provided by R01 HL80477 and R01 HL165452 from the National Heart Lung and Blood Institute (NHLBI). The authors thank the other investigators, the staff, and the participants of the REGARDS study for their valuable contributions. A full list of participating REGARDS investigators and institutions can be found at: https://www.uab.edu/soph/regardsstudy/

---

**Supplementary Figure 1:**
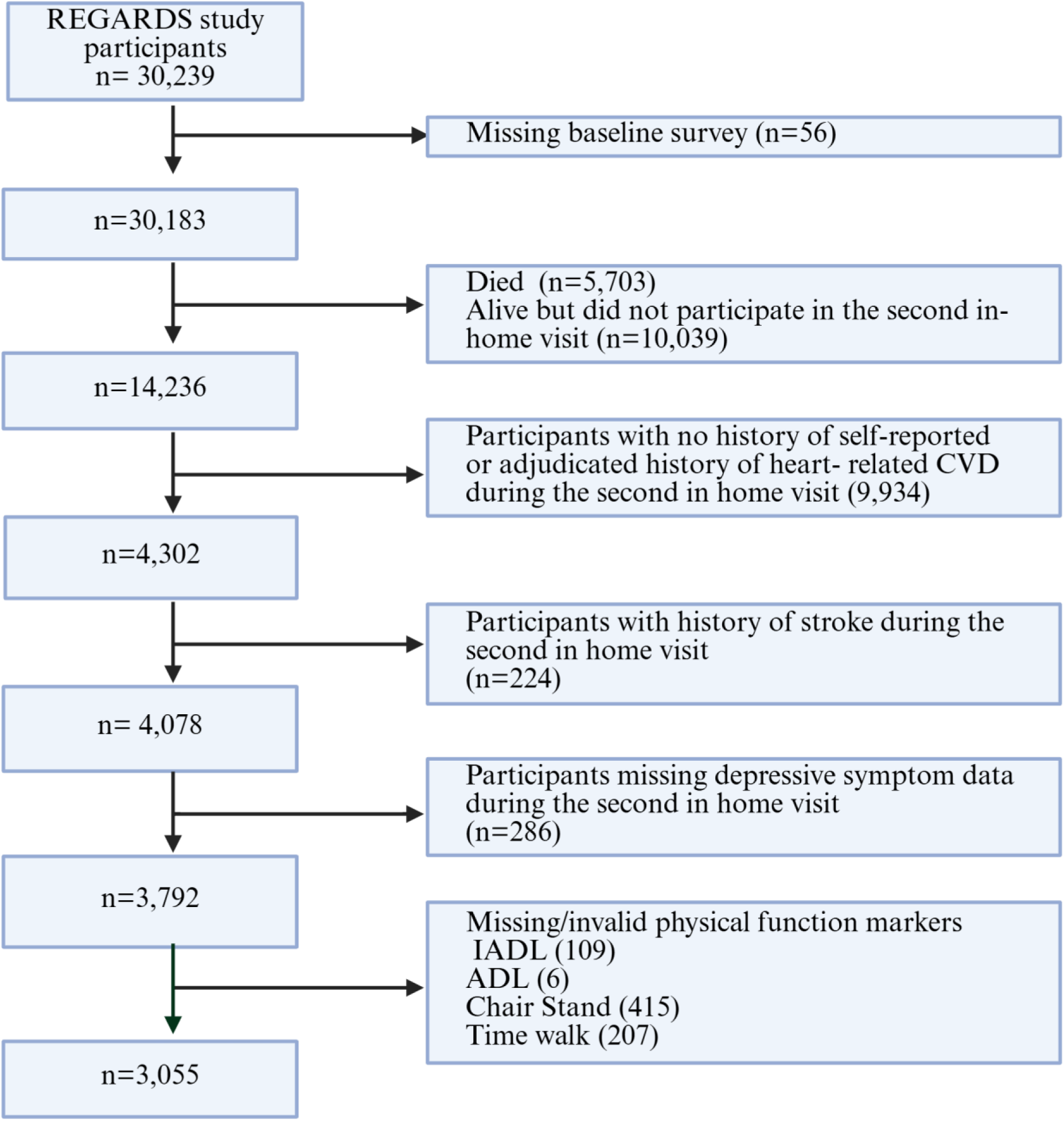
Flow diagram of REGARDS participants with heart disease included in analyses of continuous physical function outcomes.

**Supplementary Figure 2:**
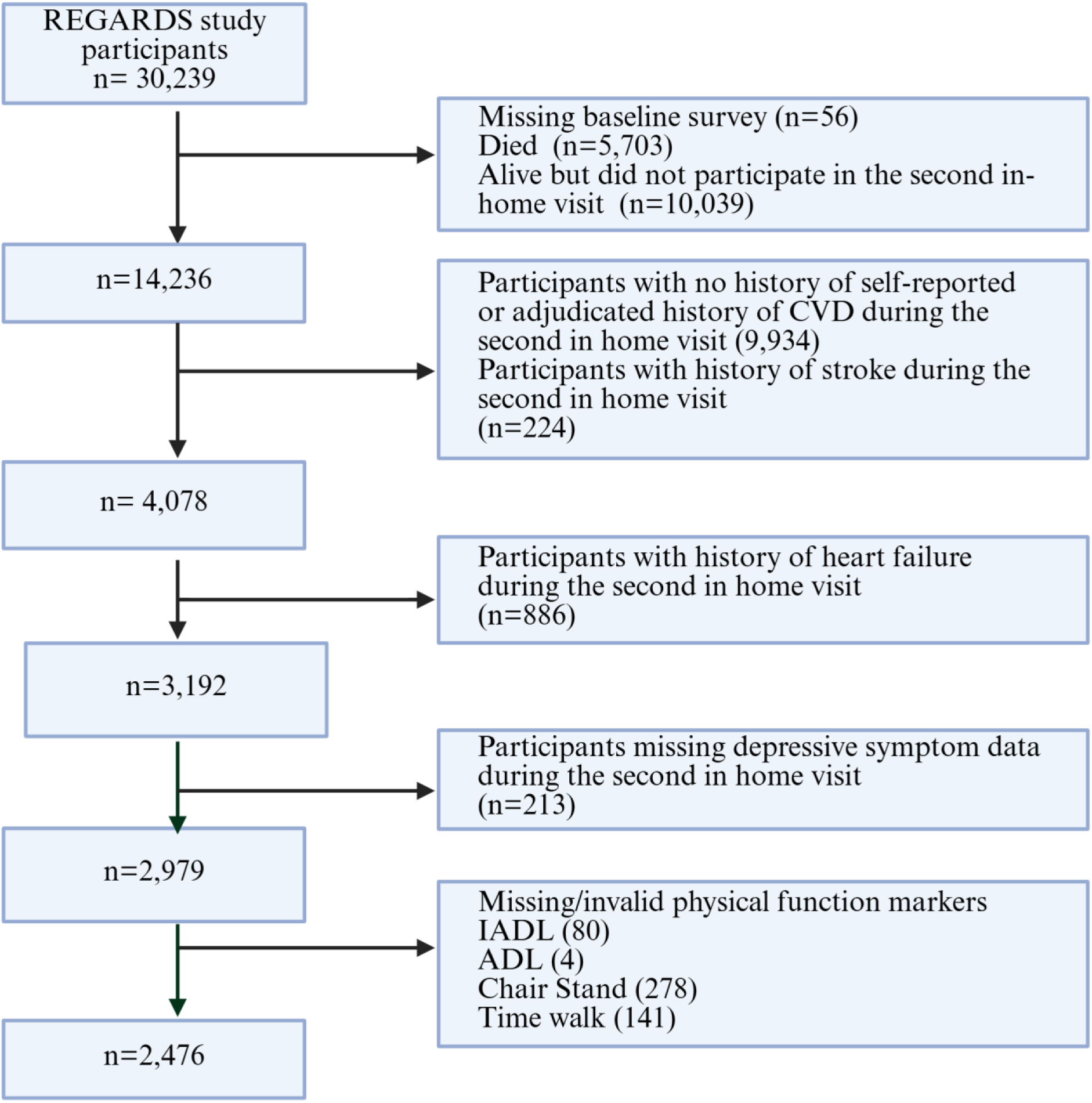
Flow diagram of REGARDS participants with heart disease excluding heart failure, for continuous physical function outcomes

**Supplementary Figure 3:**
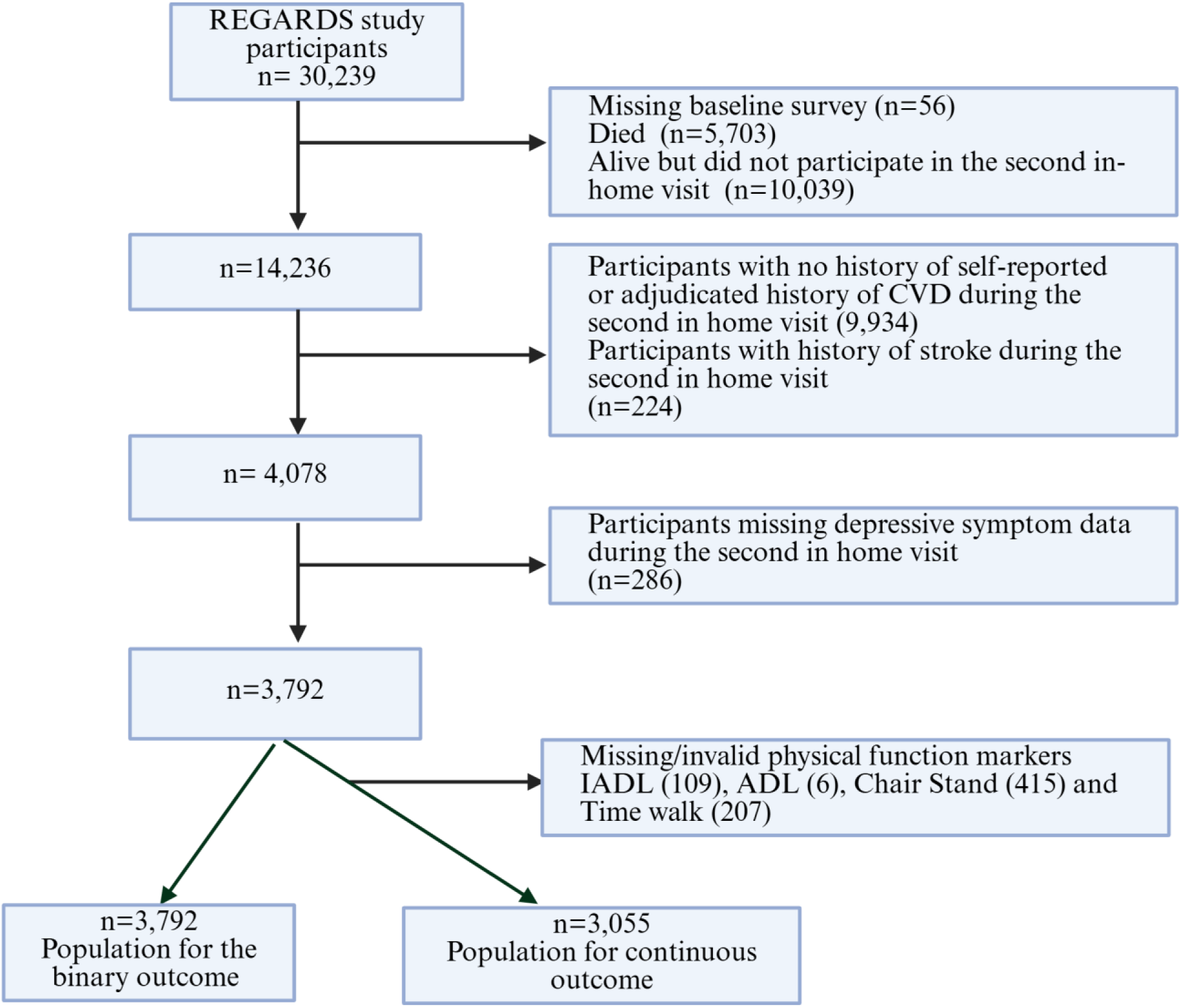
Flow diagram of REGARDS participants included in analyses of binary physical function outcomes analysis.

**Supplementary Table 1.**
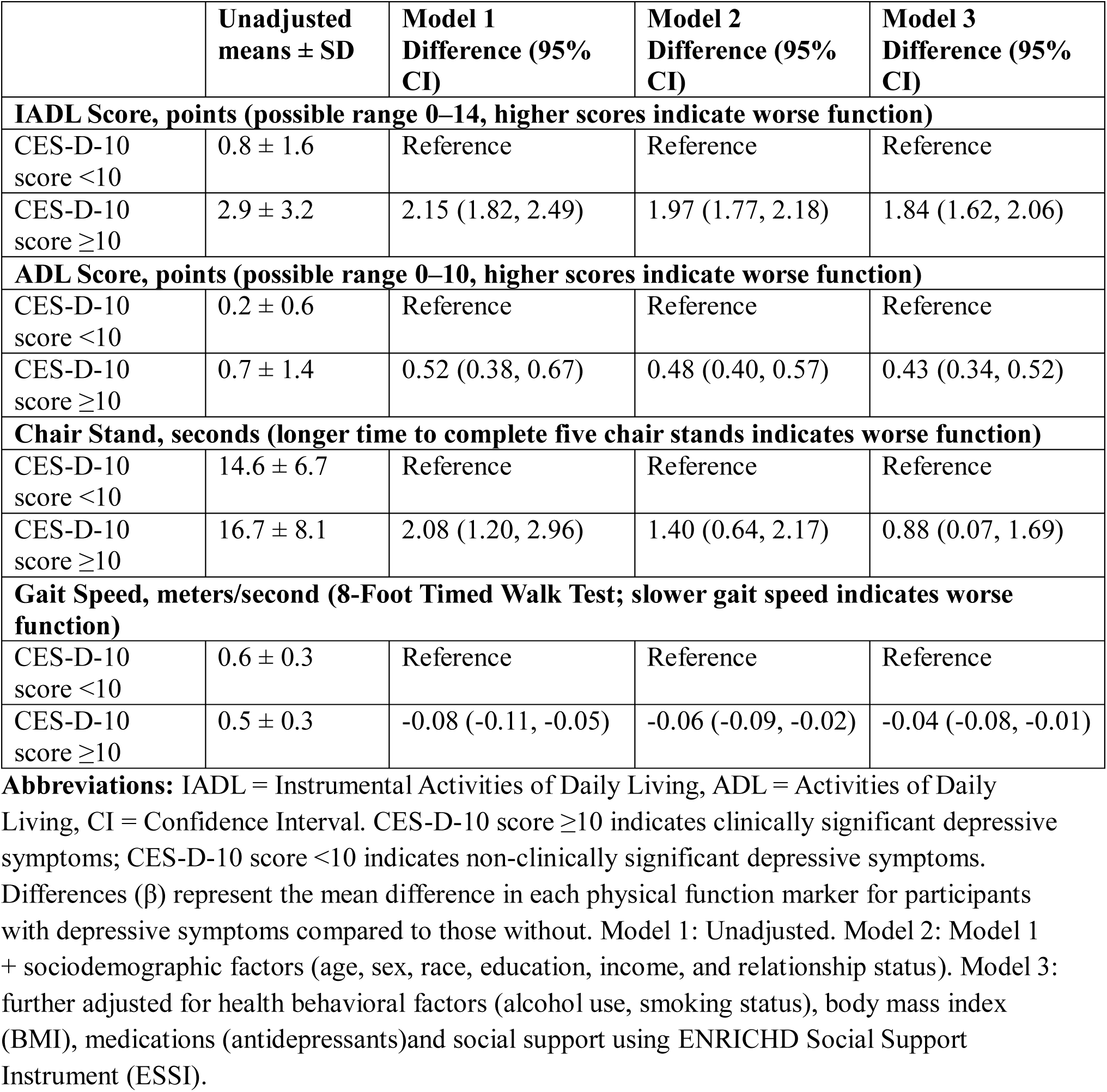
Associations between clinically significant depressive symptoms and physical function markers among REGARDS study participants with heart disease.

**Supplementary Table 2.**
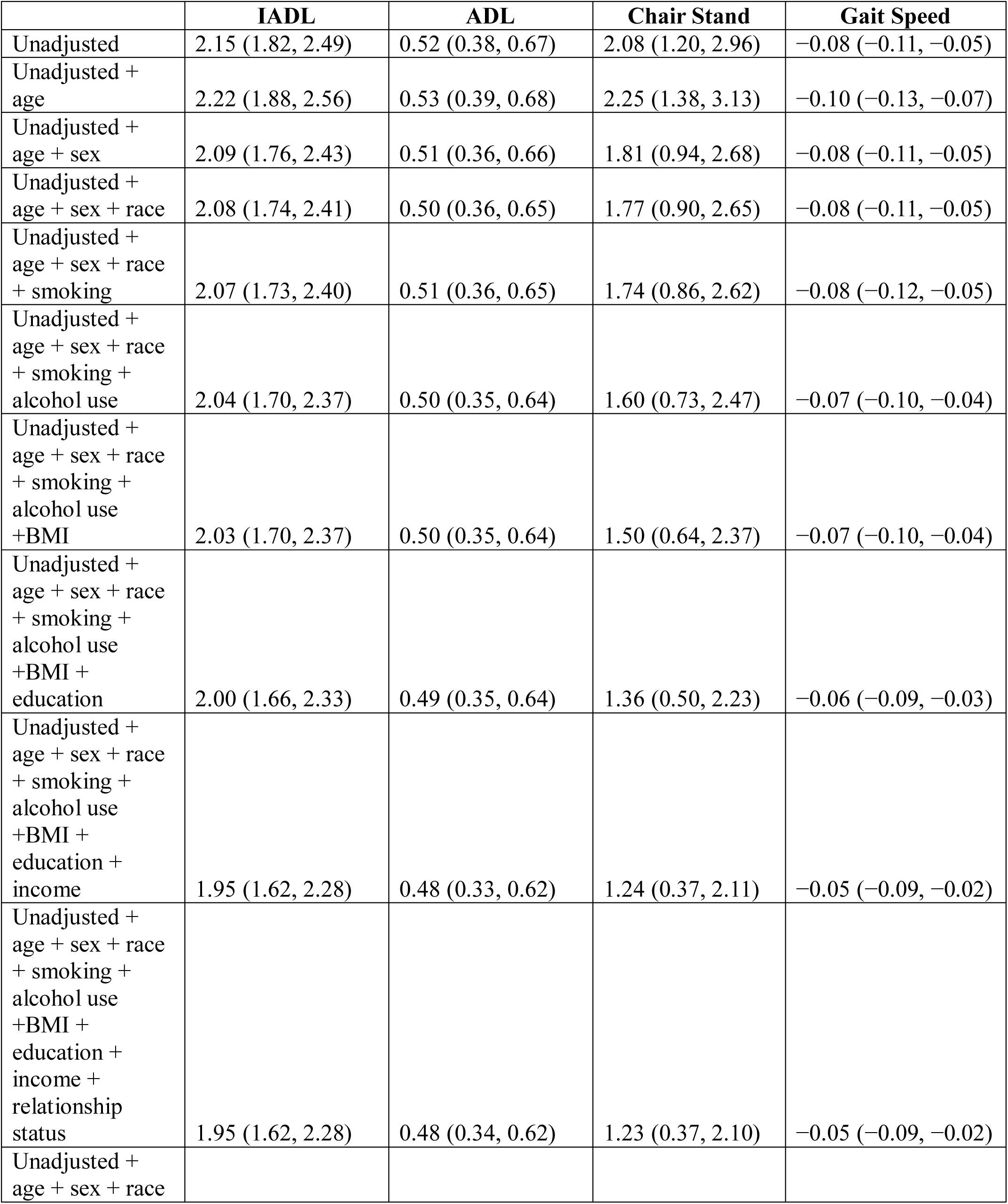
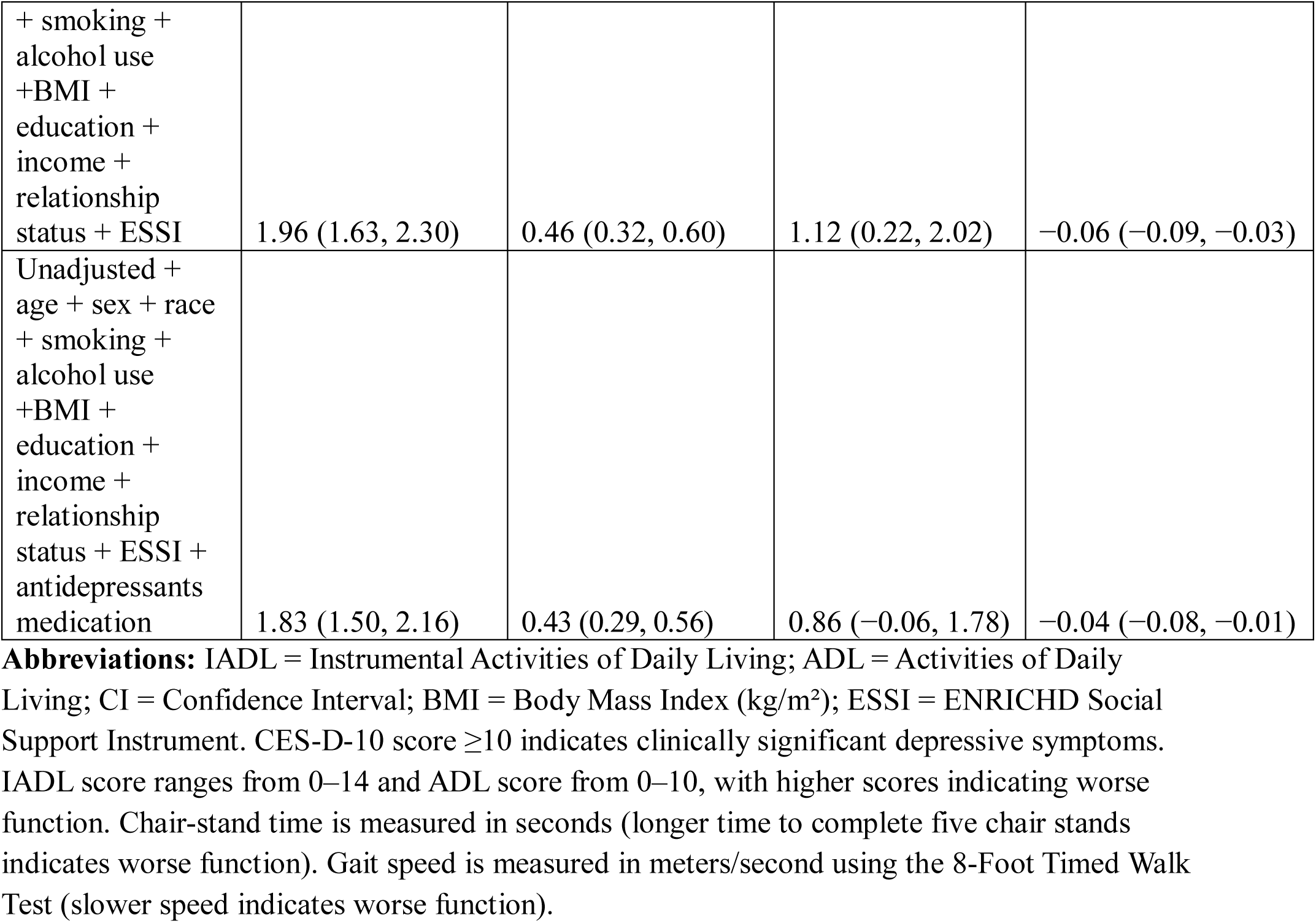
Stepwise covariate adjustment of the association between depressive symptoms and physical function outcomes.

**Supplementary Table 3.** Associations between clinically significant depressive symptoms and physical function markers excluding participants with heart failure.

| | Unadjusted means ± SD | Model 1 Difference (95% CI) | Model 2 Difference (95% CI) | Model 3 Difference (95% CI) |
| --- | --- | --- | --- | --- |
| **IADL Score, points (possible range 0–14, higher scores indicate worse function)** |  |  |  |  |
| CES-D-10 score <10 | 0.7 ± 1.5 | Reference | Reference | Reference |
| CES-D-10 score ≥10 | 3.0 ± 3.2 | 2.22 (1.85, 2.60) | 2.06 (1.84, 2.28) | 1.97 (1.74, 2.20) |
| **ADL Score, points (possible range 0–10, higher scores indicate worse function)** |  |  |  |  |
| CES-D-10 score <10 | 0.2 ± 0.6 | Reference | Reference | Reference |
| CES-D-10 score ≥10 | 0.6 ± 1.4 | 0.50 (0.34, 0.66) | 0.46 (0.37, 0.55) | 0.45 (0.35, 0.54) |
| **Chair Stand, seconds (longer time to complete five chair stands indicates worse function)** |  |  |  |  |
| CES-D-10 score <10 | 14.3 ± 6.6 | Reference | Reference | Reference |
| CES-D-10 score ≥10 | 16.5 ± 8.3 | 2.20 (1.19, 3.21) | 1.43 (0.58, 2.29) | 0.88 (–0.03, 1.78) |
| **Gait Speed, meters/second (8-Foot Timed Walk Test; slower gait speed indicates worse function)** |  |  |  |  |
| CES-D-10 score <10 | 0.6 ± 0.3 | Reference | Reference | Reference |
| CES-D-10 score ≥10 | 0.6 ± 0.3 | –0.08 (–0.12, –0.04) | –0.06 (–0.10, –0.02) | –0.04 (–0.08, 0.00) |

**Abbreviations:** IADL = Instrumental Activities of Daily Living, ADL = Activities of Daily Living, CI = Confidence Interval. CES-D-10 score ≥ 10 indicates clinically significant depressive symptoms; CES-D-10 score <10 indicates non-clinically significant depressive symptoms. Differences ( $\beta$ ) represent the mean difference in each physical function marker for participants with depressive symptoms compared to those without. Model 1: Unadjusted. Model 2: Model 1 + sociodemographic factors (age, sex, race, education, income, and relationship status). Model 3: further adjusted for health behavioral factors (alcohol use, smoking status), body mass index (BMI), medications (antidepressants) and social support using ENRICH Social Support Instrument (ESSI).
Participants with Heart Failure (HF) were excluded from the study population.

**Supplementary Table 4:** Descriptive statistics of depressive symptoms and physical function by race-sex group.

|  | **Black men** | **Black women** | **White women** | **White men** |
| --- | --- | --- | --- | --- |
| **Total (N)** | 408 | 476 | 749 | 1422 |
| CES-D-10 $\geq 10$,<br>n (%) | 36 (8.8) | 95 (20.0) | 126 (16.8) | 99 (7.0) |
| **IADL Score, points (possible range 0–14, higher scores indicate worse function)** |  |  |  |  |
| mean (SD) | 0.9 (1.8) | 2.0 (2.7) | 1.2 (2.0) | 0.8 (1.6) |
| **ADL Score, points (possible range 0–10, higher scores indicate worse function)** |  |  |  |  |
| mean (SD) | 0.2 (0.7) | 0.5 (1.3) | 0.2 (0.7) | 0.2 (0.6) |
| **Chair Stand, seconds (longer time to complete five chair stands indicates worse function)** |  |  |  |  |
| mean (SD) | 14.8 (6.2) | 16.6 (7.6) | 15.7 (7.5) | 13.9 (6.4) |
| **Gait Speed, meters/second (8-Foot Timed Walk Test; slower gait speed indicates worse function)** |  |  |  |  |
| mean (SD) | 0.6 (0.3) | 0.5 (0.3) | 0.6 (0.3) | 0.7 (0.3) |

**Abbreviations:** IADL = Instrumental Activities of Daily Living, ADL = Activities of Daily Living, %= percentage

**Supplementary Table 5.** Associations between clinically significant depressive symptoms and physical function markers among REGARDS study participants with heart disease.

| | Unadjusted means ± SD | Model 1 Difference (95% CI) | Model 2 Difference (95% CI) | Model 3 Difference (95% CI) |
| --- | --- | --- | --- | --- |
| **IADL Score, points (possible range 0–14, higher scores indicate worse function)** |  |  |  |  |
| CES-D-10 score <16 | 1.0 ± 1.8 | Reference | Reference | Reference |
| CES-D-10 score ≥16 | 3.9 ± 3.7 | 2.91 (2.24, 3.59) | 2.71 (2.36, 3.06) | 2.44 (2.07, 2.80) |
| **ADL Score, points (possible range 0–10, higher scores indicate worse function)** |  |  |  |  |
| CES-D-10 score <16 | 0.2 ± 0.7 | Reference | Reference | Reference |
| CES-D-10 score ≥16 | 1.0 ± 1.8 | 0.80 (0.48, 1.13) | 0.75 (0.60, 0.89) | 0.67 (0.52, 0.81) |
| **Chair Stand, seconds (longer time to complete five chair stands indicates worse function)** |  |  |  |  |
| CES-D-10 score <16 | 14.7 ± 6.8 | Reference | Reference | Reference |
| CES-D-10 score ≥16 | 18.0 ± 9.5 | 3.30 (1.52, 5.05) | 2.66 (1.38, 3.94) | 2.05 (0.73, 3.37) |
| **Gait Speed, meters/second (8-Foot Timed Walk Test; slower gait speed indicates worse function)** |  |  |  |  |
| CES-D-10 score <16 | 0.6 ± 0.3 | Reference | Reference | Reference |
| CES-D-10 score ≥16 | 0.5 ± 0.3 | -0.09 (-0.14, -0.04) | -0.07 (-0.13, -0.01) | -0.05 (-0.12, 0.01) |

**Abbreviations:** IADL = Instrumental Activities of Daily Living, ADL = Activities of Daily Living, CI = Confidence Interval. CES-D-10 score ≥ 16 indicates clinically significant depressive symptoms; CES-D-10 score <16 indicates non-clinically significant depressive symptoms. Differences ( $\beta$ ) represent the mean difference in each physical function marker for participants with depressive symptoms compared to those without. Model 1: Unadjusted. Model 2: Model 1 + sociodemographic factors (age, sex, race, education, income, and relationship status). Model 3: Model 2 + health behavioral factors (alcohol use, smoking status), body mass index (BMI), medications (antidepressants) and social support using ENRICH Social Support Instrument (ESSI).

**Supplementary Table 6.** Associations between clinically significant depressive symptoms and ADL outcomes by sex among REGARDS study participants with heart disease.

|  | **Male** |  | **Female** |  | **P-values** |
| --- | --- | --- | --- | --- | --- |
|  | CES-D-10 score<br>≥16 | CES-D-10<br>score <16 | CES-D-10 score<br>≥16 | CES-D-10<br>score <16 |  |
| **IADL Score, points (possible range 0–14, higher scores indicate worse function)** |  |  |  |  |  |
| Model 1<br>Difference<br>(95% CI) | 2.12 (1.10,3.15) | Reference | 3.11 (2.24, 3.97) | Reference | 0.15 |
| Model 2<br>Difference<br>(95% CI) | 2.12 (1.61,2.63) | Reference | 3.04 (2.20,3.87) | Reference | 0.01 |
| Model 3<br>Difference<br>(95% CI) | 1.89 (1.37,2.40) | Reference | 2.67 (2.14,3.20) | Reference | <0.01 |
| **ADL Score, points (possible range 0–10, higher scores indicate worse function)** |  |  |  |  |  |
| Model 1<br>Difference<br>(95% CI) | 0.53 (0.04,1.02) | Reference | 0.91 (0.48,1.33) | Reference | 0.25 |
| Model 2<br>Difference<br>(95% CI) | 0.52 (0.33,0.72) | Reference | 0.88 (0.46,1.30) | Reference | 0.01 |
| Model 3<br>Difference<br>(95% CI) | 0.44 (0.24,0.64) | Reference | 0.77 (0.55,1.00) | Reference | 0.01 |

**Abbreviations:** IADL = Instrumental Activities of Daily Living, ADL = Activities of Daily Living, CI = Confidence Interval. CES-D-10 score ≥16 indicates clinically significant depressive symptoms; CES-D-10 score <16 indicates non-clinically significant depressive symptoms. Differences (β) represent the mean difference in each physical function marker for participants with depressive symptoms compared to those without. Model 1: Unadjusted. Model 2: Model 1 + sociodemographic factors (age, sex, race, education, income, and relationship status). Model 3: Model 2 + health behavioral factors (alcohol use, smoking status), body mass index (BMI), medications (antidepressants) and social support using ENRICH Social Support Instrument (ESSI). P-values correspond to the interaction term.

**Supplementary Table 7.** Associations between clinically significant depressive symptoms and physical function as dichotomous outcomes among REGARDS study participants with heart disease.

|  | **Low physical function, n (%)** | **Model 1 Odds ratios (95% CI)** | **Model 2 Odds ratios (95% CI)** | **Model 3 Odds ratios (95% CI)** |
| --- | --- | --- | --- | --- |
| **IADL Score, points ($\geq 1$, higher scores indicate worse function)** |  |  |  |  |
| CES-D-10 score <10 | 1313 (40.2) | Reference | Reference | Reference |
| CES-D-10 score $\geq 10$ | 408 (77.3) | 5.05 (4.06, 6.30) | 4.66 (3.71, 5.93) | 4.57 (3.63, 5.75) |
| **ADL Score, points ($\geq 1$, higher scores indicate worse function)** |  |  |  |  |
| CES-D-10 score <10 | 429 (13.1) | Reference | Reference | Reference |
| CES-D-10 score $\geq 10$ | 210 (39.8) | 4.35 (3.56, 5.37) | 3.94 (3.19, 4.90) | 3.74 (3.00, 4.71) |
| **Chair Stand >13.6 sec (longer time to complete five chair stands indicates worse function)** |  |  |  |  |
| CES-D-10 score <10 | 1831 (56.1) | Reference | Reference | Reference |
| CES-D-10 score $\geq 10$ | 388 (73.5) | 2.16 (1.77, 2.66) | 1.84 (1.49, 2.29) | 1.72 (1.38, 2.14) |
| **Gait Speed <0.6 meters/second (8-Foot Timed Walk Test; slower gait speed indicates worse function)** |  |  |  |  |
| CES-D-10 score <10 | 161 (4.9) | Reference | Reference | Reference |
| CES-D-10 score $\geq 10$ | 58 (10.9) | 2.39 (1.73, 3.25) | 1.93 (1.38, 2.72) | 1.92 (1.35, 2.72) |

**Abbreviations:** IADL = Instrumental Activities of Daily Living, ADL = Activities of Daily Living, CI = Confidence Interval, % = percentages. CES-D-10 score $\geq 10$ indicates clinically significant depressive symptoms; CES-D-10 score <10 indicates non-clinically significant depressive symptoms. Model 1: Unadjusted. Model 2: Model 1 + sociodemographic factors (age, sex, race, education, income, and relationship status). Model 3: Model 2 + health behavioral factors (alcohol use, smoking status), body mass index (BMI), medications (antidepressants) and social support using ENRICH Social Support Instrument (ESSI).
Analyses performed using logistic regression models
Chair stand and gait speed poor categories include those unable to do the tests.

## References

1. Tsao CW, Aday AW, Almarzooq ZI, et al. Heart Disease and Stroke Statistics—2023 Update: A Report From the American Heart Association. Circulation. 2023;147(8):e93–e621. doi:doi:10.1161/CIR.0000000000001123

2. Lee B, Wang Y, Carlson SA, et al. National, State-Level, and County-Level Prevalence Estimates of Adults Aged ≥18 Years Self-Reporting a Lifetime Diagnosis of Depression - United States, 2020. MMWR Morb Mortal Wkly Rep. Jun 16 2023;72(24):644–650. doi:10.15585/mmwr.mm7224a1

3. Khodneva Y, Goyal P, Levitan EB, et al. Depressive Symptoms and Incident Hospitalization for Heart Failure: Findings From the REGARDS Study. J Am Heart Assoc. Apr 5 2022;11(7):e022818. doi:10.1161/jaha.121.022818

4. Alcántara C, Muntner P, Edmondson D, et al. Perfect storm: concurrent stress and depressive symptoms increase risk of myocardial infarction or death. Circ Cardiovasc Qual Outcomes. Mar 2015;8(2):146–54. doi:10.1161/circoutcomes.114.001180

5. Wiklund I, Sanne H, Vedin A, Wilhelmsson C. Psychosocial outcome one year after a first myocardial infarction. Journal of Psychosomatic Research. 1984/01/01/ 1984;28(4):309–321. 10.1016/0022-3999(84)90053-9

6. Nielsen TJ, Vestergaard M, Christensen B, Christensen KS, Larsen KK. Mental health status and risk of new cardiovascular events or death in patients with myocardial infarction: a population-based cohort study. BMJ Open. 2013;3(8):e003045. doi:10.1136/bmjopen-2013-003045

7. Rumsfeld JS, Alexander KP, Goff DC, Jr., et al. Cardiovascular health: the importance of measuring patient-reported health status: a scientific statement from the American Heart Association. Circulation. Jun 4 2013;127(22):2233–49. doi:10.1161/CIR.0b013e3182949a2e

8. Coats AJS, Forman DE, Haykowsky M, et al. Physical function and exercise training in older patients with heart failure. Nat Rev Cardiol. Sep 2017;14(9):550–559. doi:10.1038/nrcardio.2017.70

9. O’Neill D, Forman DE. The importance of physical function as a clinical outcome: Assessment and enhancement. Clin Cardiol. Feb 2020;43(2):108–117. doi:10.1002/clc.23311

10. Painter P, Stewart AL, Carey S. Physical functioning: definitions, measurement, and expectations. Adv Ren Replace Ther. Apr 1999;6(2):110–23. doi:10.1016/s1073-4449(99)70028-2

11. Kenya O, Yamaoka-Tojo M, Shinichi O, et al. A combination of depression and decreased physical function further worsens the prognosis of patients with chronic cardiovascular disease. Journal of Cardiovascular Medicine and Cardiology. 04/21 2020;7:063–071. doi:10.17352/2455-2976.000115

12. Wilcox ME, Freiheit EA, Faris P, et al. Depressive symptoms and functional decline following coronary interventions in older patients with coronary artery disease: a prospective cohort study. BMC Psychiatry. Aug 4 2016;16:277. doi:10.1186/s12888-016-0986-3

13. Investigators WCftE. Effects of Treating Depression and Low Perceived Social Support on Clinical Events After Myocardial InfarctionThe Enhancing Recovery in Coronary Heart Disease Patients (ENRICHD) Randomized Trial. JAMA. 2003;289(23):3106–3116. doi:10.1001/jama.289.23.3106

14. Orth-Gomér K, Schneiderman N, Wang H-X, Walldin C, Blom M, Jernberg T. Stress Reduction Prolongs Life in Women With Coronary Disease. Circulation: Cardiovascular Quality and Outcomes. 2009;2(1):25–32. doi:doi:10.1161/CIRCOUTCOMES.108.812859

15. Demarginalizing the Intersection of Race and Sex: A Black Feminist Critique of Antidiscrimination Doctrine, Feminist Theory and Antiracist Politics. https://chicagounbound.uchicago.edu/cgi/viewcontent.cgi?article=1052&context=uclf

16. Carbado DW, Crenshaw KW, Mays VM, Tomlinson B. INTERSECTIONALITY: Mapping the Movements of a Theory. Du Bois Rev. Fall 2013;10(2):303–312. doi:10.1017/s1742058x13000349

17. Bowleg L. The problem with the phrase women and minorities: intersectionality-an important theoretical framework for public health. Am J Public Health. Jul 2012;102(7):1267–73. doi:10.2105/ajph.2012.300750

18. Howard VJ, Cushman M, Pulley L, et al. The reasons for geographic and racial differences in stroke study: objectives and design. Neuroepidemiology. 2005;25(3):135–43. doi:10.1159/000086678

19. Levitan EB, Goyal P, Ringel JB, et al. Myocardial infarction and physical function: the REasons for Geographic And Racial Differences in Stroke prospective cohort study. BMJ Public Health. 2023;1(1)doi:10.1136/bmjph-2023-000107

20. Rizvi MR, Sharma A, Malki A, Sami W. Enhancing Cardiovascular Health and Functional Recovery in Stroke Survivors: A Randomized Controlled Trial of Stroke-Specific and Cardiac Rehabilitation Protocols for Optimized Rehabilitation. J Clin Med. Oct 18 2023;12(20)doi:10.3390/jcm12206589

21. Fu H, Si L, Guo R. What Is the Optimal Cut-Off Point of the 10-Item Center for Epidemiologic Studies Depression Scale for Screening Depression Among Chinese Individuals Aged 45 and Over? An Exploration Using Latent Profile Analysis. Front Psychiatry. 2022;13:820777. doi:10.3389/fpsyt.2022.820777

22. Andresen EM, Malmgren JA, Carter WB, Patrick DL. Screening for Depression in Well Older Adults: Evaluation of a Short Form of the CES-D. American Journal of Preventive Medicine. 1994/03/01/ 1994;10(2):77–84. 10.1016/S0749-3797(18)30622-6

23. Pashmdarfard M, Azad A. Assessment tools to evaluate Activities of Daily Living (ADL) and Instrumental Activities of Daily Living (IADL) in older adults: A systematic review. Med J Islam Repub Iran. 2020;34:33. doi:10.34171/mjiri.34.33

24. Suijker JJ, van Rijn M, Ter Riet G, Moll van Charante EP, de Rooij SE, Buurman BM. Minimal Important Change and Minimal Detectable Change in Activities of Daily Living in Community-Living Older People. J Nutr Health Aging. 2017;21(2):165–172. doi:10.1007/s12603-016-0797-8

25. Puthoff ML, Saskowski D. Reliability and responsiveness of gait speed, five times sit to stand, and hand grip strength for patients in cardiac rehabilitation. Cardiopulm Phys Ther J. Mar 2013;24(1):31–7.

26. Guralnik J, Bandeen-Roche K, Bhasin SAR, et al. Clinically Meaningful Change for Physical Performance: Perspectives of the ICFSR Task Force. J Frailty Aging. 2020;9(1):9–13. doi:10.14283/jfa.2019.33

27. Physio-pedia. Five Times Sit to Stand Test. Physio-pedia. Accessed 2/20, 2026. https://www.physio-pedia.com/Five_Times_Sit_to_Stand_Test

28. Penninx BWJH, Guralnik JM, Ferrucci L, Simonsick EM, Deeg DJH, Wallace RB. Depressive Symptoms and Physical Decline in Community-Dwelling Older Persons. JAMA. 1998;279(21):1720–1726. doi:10.1001/jama.279.21.1720

29. Moise N, Khodneva Y, Jannat-Khah DP, et al. Observational study of the differential impact of time-varying depressive symptoms on all-cause and cause-specific mortality by health status in community-dwelling adults: the REGARDS study. BMJ Open. Jan 5 2018;8(1):e017385. doi:10.1136/bmjopen-2017-017385

30. Bickl TF, F.; Brand, M. Gambling behaviour and perceived social support: The role of emotional and social support as a protective factor. Journal of Gambling Issues. 2023;(52):115–137.

31. Lett E, Asabor E, Beltrán S, Cannon AM, Arah OA. Conceptualizing, Contextualizing, and Operationalizing Race in Quantitative Health Sciences Research. Ann Fam Med. Mar-Apr 2022;20(2):157–163. doi:10.1370/afm.2792

32. Adkins-Jackson PB, Chantarat T, Bailey ZD, Ponce NA. Measuring Structural Racism: A Guide for Epidemiologists and Other Health Researchers. Am J Epidemiol. Mar 24 2022;191(4):539–547. doi:10.1093/aje/kwab239

33. Kim G, Decoster J, Huang CH, Chiriboga DA. Race/ethnicity and the factor structure of the Center for Epidemiologic Studies Depression Scale: a meta-analysis. Cultur Divers Ethnic Minor Psychol. Oct 2011;17(4):381–96. doi:10.1037/a0025434

34. Boutin-Foster C. An item-level analysis of the Center for Epidemiologic Studies Depression Scale (CES-D) by race and ethnicity in patients with coronary artery disease. Int J Geriatr Psychiatry. Oct 2008;23(10):1034–9. doi:10.1002/gps.2029

35. Burrows BT, Sloane R, Ashner MC, et al. Associations of depressive symptoms and antidepressant medication use with physical function among middle-aged adults: Results from the CARDIA Function study. J Affect Disord. May 15 2026;401:121324. doi:10.1016/j.jad.2026.121324

36. Li X, Zhou J, Wang M, Yang C, Sun G. Cardiovascular disease and depression: a narrative review. Front Cardiovasc Med. 2023;10:1274595. doi:10.3389/fcvm.2023.1274595

37. Björgvinsson T, Kertz SJ, Bigda-Peyton JS, McCoy KL, Aderka IM. Psychometric Properties of the CES-D-10 in a Psychiatric Sample. Assessment. 2013;20(4):429–436. doi:10.1177/1073191113481998

38. Williams MW, Li CY, Hay CC. Validation of the 10-item Center for Epidemiologic Studies Depression Scale Post Stroke. J Stroke Cerebrovasc Dis. Dec 2020;29(12):105334. doi:10.1016/j.jstrokecerebrovasdis.2020.105334

39. Guralnik JM, Simonsick EM, Ferrucci L, et al. A short physical performance battery assessing lower extremity function: association with self-reported disability and prediction of mortality and nursing home admission. J Gerontol. Mar 1994;49(2):M85–94. doi:10.1093/geronj/49.2.m85

40. Peel NM, Kuys SS, Klein K. Gait Speed as a Measure in Geriatric Assessment in Clinical Settings: A Systematic Review. The Journals of Gerontology: Series A. 2012;68(1):39–46. doi:10.1093/gerona/gls174

41. Physiopedia. Gait Speed as an Objective Measure. https://www.physio-pedia.com/Gait_Speed_as_an_Objective_Measure

42. Lenze EJ, Schulz R, Martire LM, et al. The Course of Functional Decline in Older People with Persistently Elevated Depressive Symptoms: Longitudinal Findings from the Cardiovascular Health Study. Journal of the American Geriatrics Society. 2005;53(4):569–575. 10.1111/j.1532-5415.2005.53202.x

43. Wand BM, Chiffelle LA, O’Connell NE, McAuley JH, Desouza LH. Self-reported assessment of disability and performance-based assessment of disability are influenced by different patient characteristics in acute low back pain. Eur Spine J. Apr 2010;19(4):633–40. doi:10.1007/s00586-009-1180-9

44. Mastenbroek MH, Versteeg H, Zijlstra WP, Meine M, Spertus JA, Pedersen SS. Disease-specific health status as a predictor of mortality in patients with heart failure: a systematic literature review and meta-analysis of prospective cohort studies. European Journal of Heart Failure. 2014;16(4):384–393. 10.1002/ejhf.55

45. Xu L, Zhai X, Shi D, Zhang Y. Depression and coronary heart disease: mechanisms, interventions, and treatments. Review. Frontiers in Psychiatry. 2024-February-09 2024;Volume 15 - 2024doi:10.3389/fpsyt.2024.1328048

46. Choi NG, Kim J, Marti CN, Chen GJ. Late-life depression and cardiovascular disease burden: examination of reciprocal relationship. Am J Geriatr Psychiatry. Dec 2014;22(12):1522–9. doi:10.1016/j.jagp.2014.04.004

47. Schulz R, Beach SR, Ives DG, Martire LM, Ariyo AA, Kop WJ. Association Between Depression and Mortality in Older Adults: The Cardiovascular Health Study. Archives of Internal Medicine. 2000;160(12):1761–1768. doi:10.1001/archinte.160.12.1761

48. Bigger JT, Glassman AH. The American Heart Association science advisory on depression and coronary heart disease: an exploration of the issues raised. Cleve Clin J Med. Jul 2010;77 Suppl 3:S12–9. doi:10.3949/ccjm.77.s3.03

49. Huffman JC, Adams CN, Celano CM. Collaborative Care and Related Interventions in Patients With Heart Disease: An Update and New Directions. Psychosomatics. Jan-Feb 2018;59(1):1–18. doi:10.1016/j.psym.2017.09.003

50. van den Houdt SCM, Mommersteeg PMC, Widdershoven J, Kupper N. Sex and Gender Differences in Psychosocial Risk Profiles Among Patients with Coronary Heart Disease - the THORESCI-Gender Study. Int J Behav Med. Feb 2024;31(1):130–144. doi:10.1007/s12529-023-10170-5

51. Orth-Gomér K, Schneiderman N, Wang HX, Walldin C, Blom M, Jernberg T. Stress reduction prolongs life in women with coronary disease: the Stockholm Women’s Intervention Trial for Coronary Heart Disease (SWITCHD). Circ Cardiovasc Qual Outcomes. Jan 2009;2(1):25–32. doi:10.1161/circoutcomes.108.812859

52. Crenshaw KW. Demarginalizing the intersection of race and sex: A Black feminist critique of antidiscrimination doctrine, feminist theory and antiracist politics. University of Chicago Legal Forum. 1989;1989(1):139–167.

53. Pinto Pereira SM, Geoffroy M-C, Power C. Depressive Symptoms and Physical Activity During 3 Decades in Adult Life: Bidirectional Associations in a Prospective Cohort Study. JAMA Psychiatry. 2014;71(12):1373–1380. doi:10.1001/jamapsychiatry.2014.1240

54. Besser LM, Brenowitz WD, Meyer OL, Hoermann S, Renne J. Methods to Address Self-Selection and Reverse Causation in Studies of Neighborhood Environments and Brain Health. Int J Environ Res Public Health. Jun 16 2021;18(12)doi:10.3390/ijerph18126484

